# Evaluating the Large Language Model-Based Quality Assurance Tool for Auto-Contouring

**DOI:** 10.64898/2026.03.31.26349802

**Authors:** Ryota Tozuka, Tomoko Akita, Masaki Matsuda, Hikaru Tanno, Masahide Saito, Hikaru Nemoto, Koji Mitsuda, Noriyuki Kadoya, Keiichi Jingu, Hiroshi Onishi

## Abstract

**Purpose:** Manual verification of AI-based auto-contouring is labor-intensive and prone to fatigue-related errors. This study developed a large language model (LLM)-based automated Quality Assurance (QA) for auto-contouring (LAQUA) system using a multimodal LLM, Gemini 2.5 Pro, and evaluated its feasibility as a clinical primary screening tool to streamline the QA workflow.

**Methods:** Twenty male pelvic CT scans from an open dataset were utilized to generate auto-contours of the bladder, prostate, rectum, and bilateral femoral heads using three distinct software packages (OncoStudio, RatoGuide prototype, and syngo.via). The generated contours for each slice were exported as PDF images with overlaid contours and input into Gemini 2.5 Pro. The LLM was instructed to rate the contour quality on a 5-point clinical scale (5: Optimal; 4: Acceptable; 3: Suboptimal; 2: Unacceptable; redraw from scratch; 1: Unacceptable; organ not detected or completely wrong). Spearman’s rank correlation coefficients (ρ) and weighted kappa coefficients (κ) were calculated based on evaluations by two board-certified radiation oncologists as ground truth. Additionally, to assess screening performance, sensitivity and specificity were calculated by dichotomizing the scores, defining “inadequate” contours (scores < 3 or < 4) as the target for detection, compared to “adequate” contours (scores ≥ 3 or ≥ 4). Finally, the alignment of the rationales provided by the LLM with the auto-contouring quality was evaluated by the same two board-certified radiation oncologists. This was conducted using a Likert scale assessing four domains (error detection, hallucination, clinical relevance, and anatomical understanding), each scored out of 2 points.

**Results:** The LAQUA system demonstrated moderate to strong agreement with expert judgments across all evaluated software (ρ: 0.733–0.794; quadratic weighted κ: 0.730–0.798) and organs (ρ: 0.567–0.835; quadratic weighted κ: 0.639–0.804). Regarding screening performance, a cutoff of ≥3 as “adequate” achieved the highest sensitivity and specificity in specific subgroups, but with wide 95% confidence intervals (CIs). A cutoff of ≥4 as “adequate “ narrowed the CIs, yielding the highest sensitivity in the rectum (0.976) and the highest specificity in the left femoral head (0.933). Qualitatively, the LLM’s rationales achieved an overall mean score of 1.70 ± 0.48 (out of 2), with 155 of 291 outputs receiving perfect scores across all criteria.

**Conclusions:** The LAQUA system demonstrated substantial agreement with expert evaluations in AI-based auto-contouring quality assessment. While potential overestimation bias (risk of missing “inadequate” cases) warrants caution, the observed sensitivity suggests its feasibility as a primary screening QA tool to efficiently filter acceptable contours, thereby reducing the clinical workload.

## Introduction

In recent years, the integration of artificial intelligence (AI)-based auto-contouring (AC) technologies has been rapidly advancing in clinical radiotherapy^1–3^. These technologies have been shown not only to significantly reduce the time-consuming labor of manual contouring during treatment planning but also to minimize inter-operator variability, thereby contributing to the standardization of treatment quality^4,5^. However, current AC systems do not always guarantee accurate results. Instances of inaccurate contour outputs have been reported due to image artifacts, individual anatomical variations, and biases in training data^6,7^. If these inaccuracies are overlooked and applied clinically without correction, they can lead to the overestimation or underestimation of dose evaluations for target volumes and organs at risk (OARs). This poses a severe risk to patient prognosis and safety, potentially resulting in adverse events. Therefore, it is of paramount importance to conduct robust quality assurance (QA) for AC^8^.

At present, QA of AC is predominantly performed through the visual inspection of generated contours by experts such as radiation oncologists and medical physicists. However, the manual review of numerous contours across hundreds of slices is highly time-consuming and inherently carries the risk of oversight. In particular, AC can fail even in the absence of obvious confounding factors^9,10^. Such unexpected errors can induce automation bias in evaluators, thereby elevating the risk that critical mistakes will be overlooked^11,12^.

To address this issue, research has been conducted on applying AI technologies to automate the QA of AC. Several previous studies have performed validation by calculating geometric metrics between contours generated by the primary AC system and those from independent AC systems^13,14^. Although these models demonstrated favorable predictive accuracy, it has frequently been pointed out that geometric metrics do not necessarily correlate with clinical assessments; therefore, relying solely on these metrics makes clinical implementation difficult. Furthermore, while other studies have reported attempts to classify AC outcomes as “pass/fail” or to detect failures based on contour features, these approaches were limited to merely outputting judgment results^15^. In a recent study, Luan et al. proposed a system that attempts to classify and automatically correct contouring errors using multi-modal vision-language representations^16^. However, the utilization of language information in this approach is limited to extracting text features by fitting predicted error classifications into predefined, fixed templates. Consequently, these approaches remain confined to merely outputting judgment results or boilerplate text. They fall short of flexibly describing complex error situations in natural language for experts, and they fail to provide detailed, context-specific explanations of exactly which regions of the contour require what type of modification.

Large Language Models (LLMs) represent one of the most actively researched topics today. Recent evidence has demonstrated that these models possess a profound understanding of the medical domain, particularly in the field of radiology^17–19^. We hypothesized that by leveraging the advanced visual perception and exceptional linguistic generation capabilities of modern LLMs, it would be possible to pinpoint specific AC failures and describe their nature using natural language. Based on this hypothesis, this study developed and evaluated the LLM-based automated quality assurance for auto-contouring (LAQUA) system, designed to perform fully automated QA for AC through the application of LLMs.

## Materials and methods

### Patient Data

In this study, a total of 20 male pelvic cases were selected from a publicly available dataset established in a previous study^20^. The target structures for this study were the bladder, prostate, rectum, and bilateral femoral heads. Specifically, the initial selection comprised cases with IDs ranging from 1 to 22. From this cohort, two cases were excluded: ID 9, due to missing rectal contour data, and ID 21, as the data file itself was unavailable. A notable characteristic of this dataset is the inclusion of anatomical edge cases. This composition enables a comprehensive evaluation of the LAQUA system’s robustness across not only typical but also atypical cases. Detailed information regarding the data collection protocols and imaging parameters can be found in the original publication^20^.

### Auto-Contour Generation

In this study, three different commercial software platforms were employed for AC generation: OncoStudio 2.0.4 (Oncosoft Inc., Seoul, Republic of Korea), RatoGuide 1.2.6 prototype (AiRato Inc., Sendai, Japan), and syngo.via VB60A_HF04 (Siemens Healthineers, Forchheim, Germany)^10,21,22^. The data input process for each software was conducted using a standardized procedure. First, the CT datasets of the 20 selected cases were imported into each software. Subsequently, AC was performed using the default settings of each platform. The regions of interest for evaluation were defined as the following five organs, for which ground truth contours were provided in the public dataset: the prostate, bladder, rectum, and left and right femoral heads.

### Construction of the LAQUA System

An overview of the LAQUA system is illustrated in Figure 1. The entire pipeline, encompassing image processing and the LLM inference process, was fully automated using Python 3.12. To prepare visual input data for the LLM while preserving three-dimensional continuity, the organ contours generated by AC were overlaid as red lines on the CT images and converted into a PDF format (one slice per page for each organ) using the matplotlib library in Python. To ensure optimal visibility of soft tissues, standard image window settings were applied with a Window Level of 40 HU and a Window Width of 350 HU. To allow the LLM to comprehend the relative spatial relationships between the target and surrounding OARs or anatomical structures, image cropping was not performed, preserving the full field of view of the CT images. Furthermore, to enable the recognition of craniocaudal boundaries, a margin of three additional slices extending craniocaudally beyond the AC-delineated extent was included, thereby facilitating the evaluation of craniocaudal adequacy.

**Figure 1.**
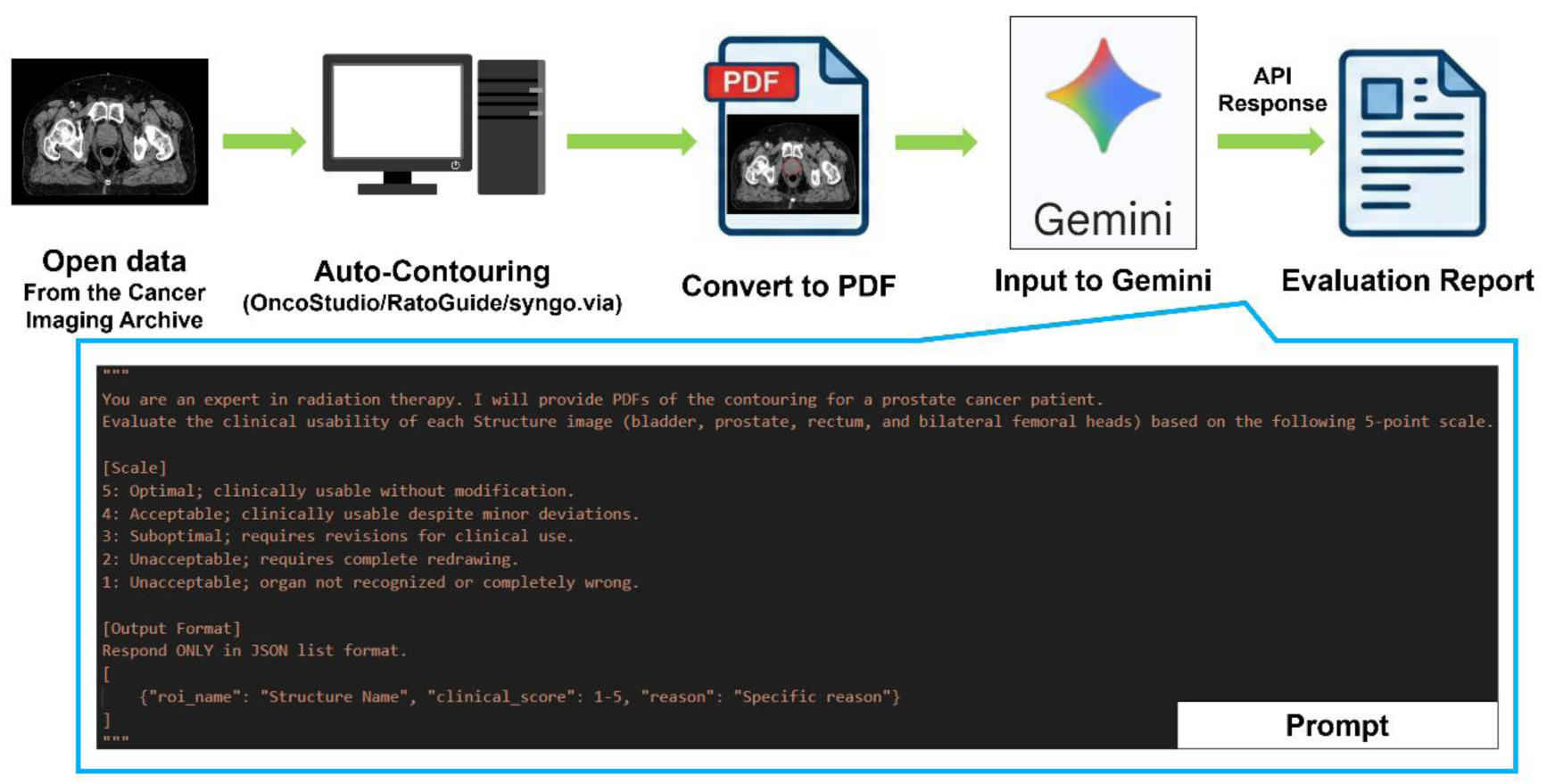
Workflow of the proposed system.

Regarding the hyperparameters of the LLM, setting the temperature as close to 0 as possible is desirable to ensure output reproducibility. However, Google has reported that setting it strictly to 0 can destabilize the model’s output, potentially leading to infinite generation loops. Therefore, based on Google’s recommended settings, the temperature was set to 0.1 in this study^23^.

Gemini 2.5 Pro, a multimodal large language model, was adopted as the evaluation model and executed via the Gemini API. A maximum of five PDFs (prostate, bladder, rectum, and left and right femoral heads) were generated per patient and subsequently fed into the LLM on a per-patient basis. The prompt used to execute the evaluation task was as follows:

*You are an expert in radiation therapy. I will provide PDFs of the contouring for a prostate cancer patient*.

*Evaluate the clinical usability of each Structure image (bladder, prostate, rectum, and bilateral femoral heads) based on the following 5-point scale*.

*[Scale]*

*5: Optimal; clinically usable without modification*.

*4: Acceptable; clinically usable despite minor deviations. 3: Suboptimal; requires revisions for clinical use*.

*2: Unacceptable; requires complete redrawing*.

*1: Unacceptable; organ not recognized or completely wrong*.

*[Output Format]*

*Respond ONLY in JSON list format. [*

*{“roi_name”: “Structure Name”, “clinical_score”: 1-5, “reason”: “Specific reason”}*

]

The LLM was instructed to generate structured output based on the aforementioned prompt. First, it was tasked with scoring the quality of the AC on a 5-point Likert scale. The evaluation criteria are detailed in Table 1. Second, it was required to provide a natural language description explaining the rationale behind its judgment. This represents the most significant advantage of utilizing an LLM, as it enables the provision of human-interpretable information regarding anatomical structures and contour deviations, rather than merely outputting simple numerical values.

**Table 1.**
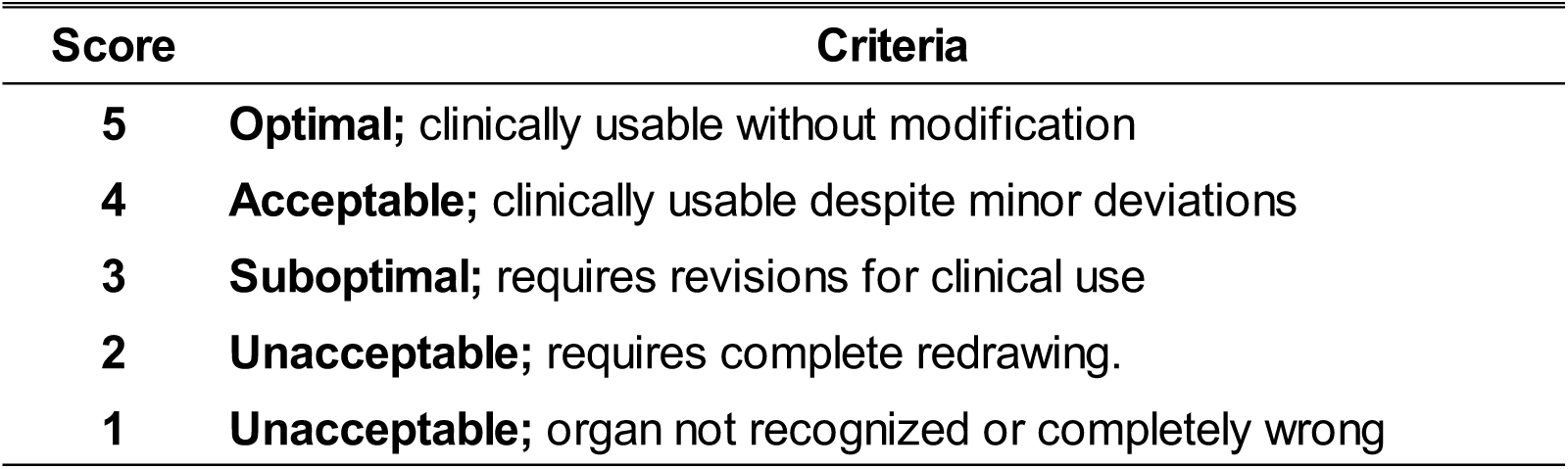
Qualitative evaluation criteria for auto-contours.

### Evaluation

The evaluation process comprised three stages: verification of the geometric accuracy of the AC, evaluation of the LLM-derived quality scores, and validation of the rationales underlying the LLM’s judgments.

In the first stage, to grasp the overall quality trends of the auto-contours, an evaluation using geometric metrics was conducted. Using the contours provided in the public dataset as the ground truth, the following three metrics were calculated: volume Dice Similarity Coefficient (vDSC), 95th percentile Hausdorff Distance (HD95), and surface Dice Similarity Coefficient (sDSC)^24^. For the sDSC, the acceptable tolerance threshold 𝜏 was set to 1 mm in this study^5,25,26^.

In the second stage, the concordance between the 5-point AC quality scores output by the LLM and the evaluation scores provided by human experts was evaluated. Two radiation oncologists with over 10 years of clinical experience served as human evaluators and assessed the AC quality by consensus, applying the same criteria used by the LLM (Table 1). To evaluate the correlation between the two sets of scores, Spearman’s rank correlation coefficient was calculated. Additionally, the quadratic weighted kappa coefficient was computed as a metric that accounts for the magnitude of score discrepancies. Furthermore, to verify the utility of the LLM as a clinical screening tool, the 5-point scores were dichotomized into “adequate” (scores ≥ 3 or ≥ 4) and “inadequate” (scores < 3 or < 4) categories based on a predefined cutoff, and the sensitivity and specificity for detecting inadequate ACs were calculated.

In the third stage, the rationales provided by Gemini 2.5 Pro for its judgments were evaluated for their alignment with the actual AC quality. Specifically, the same two radiation oncologists from the second stage visually assessed whether the error locations and rationales identified by the LLM matched the true state of the auto-contours, utilizing a novel Likert scale detailed in Table 2.

**Table 2.**
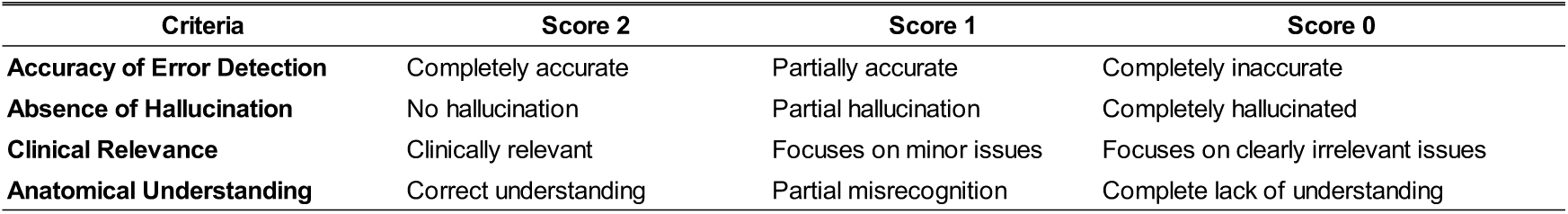
Scoring criteria for the visual evaluation of large language model performance.

| Criteria | Score 2 | Score 1 | Score 0 |
| --- | --- | --- | --- |
| Accuracy of Error Detection | Completely accurate | Partially accurate | Completely inaccurate |
| Absence of Hallucination | No hallucination | Partial hallucination | Completely hallucinated |
| Clinical Relevance | Clinically relevant | Focuses on minor issues | Focuses on clearly irrelevant issues |
| Anatomical Understanding | Correct understanding | Partial misrecognition | Complete lack of understanding |

## Result

### Quantitative Evaluation using Geometric Metrics

Figure 2 shows the results of the quantitative evaluation using geometric metrics to verify the baseline quality of the evaluated AC software. The mean vDSC values were 0.89 ± 0.11 (OncoStudio), 0.82 ± 0.19 (RatoGuide), and 0.86 ± 0.14 (syngo.via). Overall, all three software platforms demonstrated good geometric agreement (Dice coefficient ≥ 0.8)^27,28^. However, numerous instances of contours with large errors, which were plotted as outliers originating from atypical cases, were also observed. In particular, the delineation accuracy for the prostate tended to be relatively lower than that for OARs. For HD95 and sDSC, which are less susceptible to volume effects, the values were 5.25 ± 5.56 and 0.64 ± 0.23 (OncoStudio), 7.44 ± 7.90 and 0.57 ± 0.29 (RatoGuide), and 6.38 ± 6.25 and 0.56 ± 0.20 (syngo.via), respectively.

**Figure 2.**
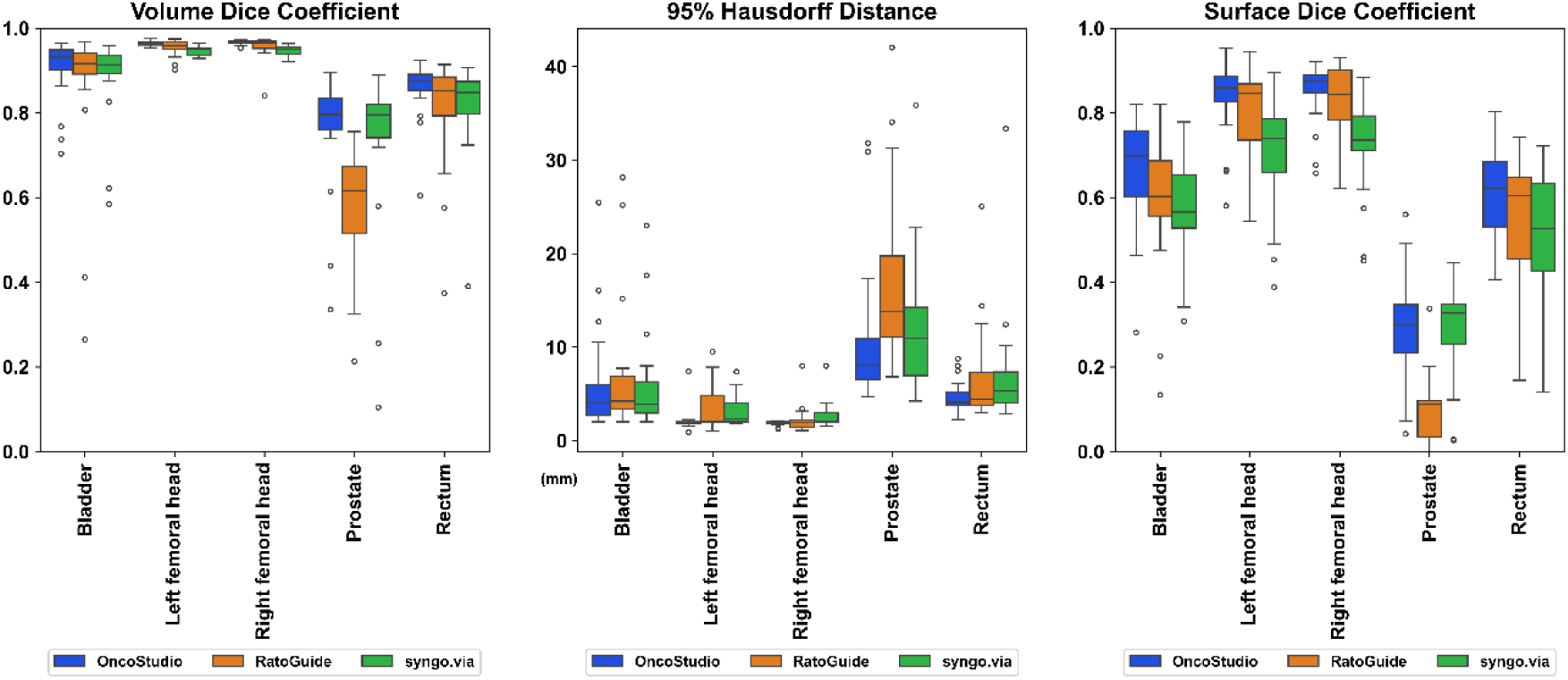
Box plots of volume Dice Similarity Coefficient, 95th percentile Hausdorff Distance, and surface Dice Similarity Coefficient for the three auto-contouring software platforms. The horizontal line within each box represents the median, and the whiskers extend to 1.5 times the interquartile range. Open circles represent outliers.

### Concordance Between LLM and Expert Evaluations

Figure 3 shows the confusion matrices comparing the evaluation scores assigned by the LLM with those given by the radiation oncologist, categorized by software and organ, respectively. Overall, the LLM evaluations exhibited a strong correlation with the radiation oncologist’s judgments. Spearman’s rank correlation coefficients and the quadratic weighted kappa coefficients were 0.733 and 0.730 for OncoStudio, 0.794 and 0.765 for RatoGuide, and 0.790 and 0.798 for syngo.via, respectively. Both metrics indicated strong or substantial agreement^29,30^. Furthermore, organ-specific analyses demonstrated moderate or higher correlations across all evaluated structures, with the detailed results summarized in Table 3. The highest agreement was observed in the rectum (Spearman’s rank correlation coefficient = 0.835, quadratic weighted kappa = 0.804), whereas the left femoral head exhibited the lowest correlation (Spearman’s rank correlation coefficient = 0.567, quadratic weighted kappa = 0.639).

**Figure 3.**
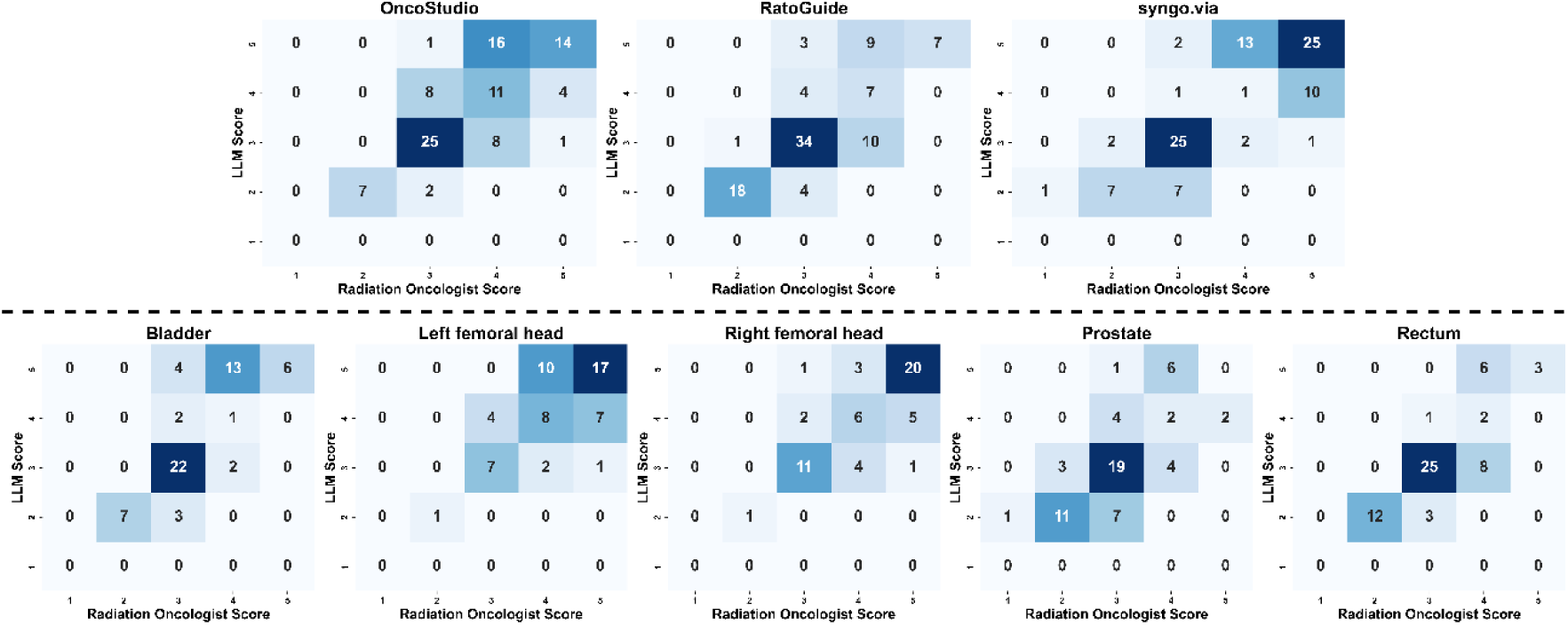
Confusion matrices comparing the LLM scores (y-axis) and the radiation oncologist scores (x-axis) for the auto-contouring results. The top row shows the results across the three auto-contouring software platforms, while the bottom row displays the results for each of the five organs. The numbers within the cells represent the number of cases, with darker colors indicating higher frequencies. LLM: Large Language Model

**Table 3.**
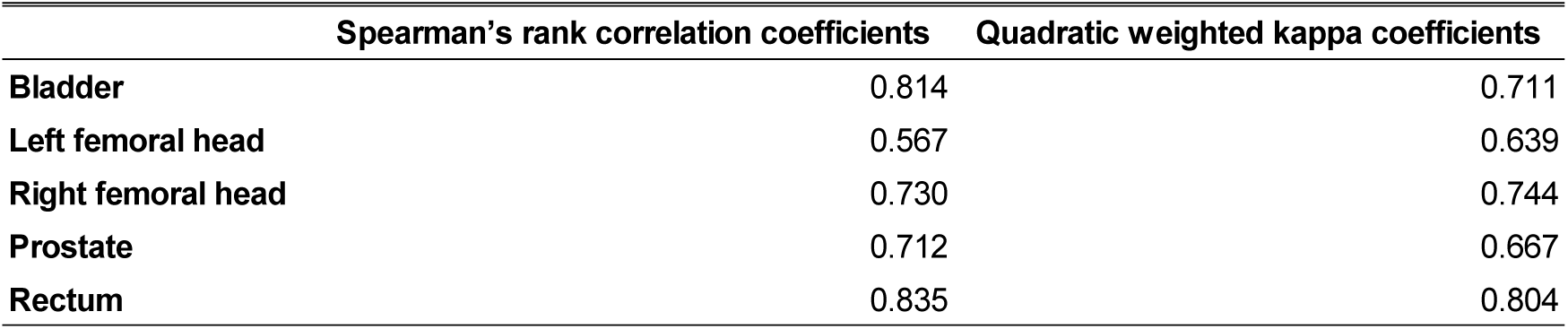
Correlation between large language model evaluations and radiation oncologist judgments by organ.

Next, regarding acceptable criteria for clinical use, two conditions were established: defining a score of ≥ 3 as “adequate,” and defining a score of ≥ 4 as “adequate.” Tables 4 and 5 summarize the sensitivity and specificity for detecting inadequate auto-contours when dichotomized based on these criteria, categorized by software and by organ, respectively. When a score of ≥ 3 was defined as “adequate” (i.e., assessing whether redrawing from scratch is necessary), the software exhibiting the highest sensitivity and specificity was OncoStudio (1.000 and 0.978, respectively). In the organ-specific analysis, bilateral femoral heads achieved the highest performance (sensitivity: 1.000, specificity: 1.000). However, wide 95% confidence intervals (CIs) were observed, indicating residual statistical uncertainty. When a score of ≥ 4 was defined as “adequate” (i.e., assessing whether the AC is usable but requires modification), the software exhibiting the highest sensitivity and specificity was syngo.via (sensitivity: 0.933, specificity: 0.942). In the organ-specific analysis, the highest sensitivity was observed in the rectum (0.976), whereas the highest specificity was observed in the left femoral head (0.933). Although the sensitivities slightly decreased compared to the condition where a score of ≥ 3 was defined as “adequate,” the widths of the 95% CIs narrowed (e.g., from 0.490–0.943 to 0.821–0.977 for syngo.via).

**Table 4.** Sensitivity, specificity, and accuracy (with 95% confidence intervals) for detecting inadequate automated contours by software.

| Defining a score of $\geq 3$ as "adequate" | Sensitivity | Specificity | Accuracy |
| --- | --- | --- | --- |
| OncoStudio | 1.000 (0.646–1.000) | 0.978 (0.923–0.994) | 0.979 (0.928–0.994) |
| RatoGuide | 0.947 (0.754–0.991) | 0.949 (0.875–0.980) | 0.948 (0.885–0.978) |
| syngo.via | 0.800 (0.490–0.943) | 0.920 (0.843–0.960) | 0.907 (0.833–0.950) |

| Defining a score of $\geq 4$ as "adequate" | Sensitivity | Specificity | Accuracy |
| --- | --- | --- | --- |
| OncoStudio | 0.791 (0.648–0.886) | 0.833 (0.713–0.910) | 0.814 (0.726–0.879) |
| RatoGuide | 0.891 (0.791–0.946) | 0.697 (0.527–0.826) | 0.825 (0.737–0.888) |
| syngo.via | 0.933 (0.821–0.977) | 0.942 (0.844–0.980) | 0.938 (0.872–0.971) |

**Table 5.** Sensitivity, specificity, and accuracy (with 95% confidence intervals) for detecting inadequate automated contours by organ.

| Defining a score of $\geq 3$ as "adequate" | Sensitivity | Specificity | Accuracy |
| --- | --- | --- | --- |
| Bladder | 1.000 (0.646–1.000) | 0.943 (0.846–0.981) | 0.950 (0.863–0.983) |
| Left femoral head | 1.000 (0.207–1.000) | 1.000 (0.936–1.000) | 1.000 (0.937–1.000) |
| Right femoral head | 1.000 (0.207–1.000) | 1.000 (0.932–1.000) | 1.000 (0.934–1.000) |
| Prostate | 0.800 (0.548–0.930) | 0.844 (0.712–0.923) | 0.833 (0.720–0.907) |
| Rectum | 1.000 (0.758–1.000) | 0.938 (0.832–0.979) | 0.950 (0.863–0.983) |

| Defining a score of $\geq 4$ as "adequate" | Sensitivity | Specificity | Accuracy |
| --- | --- | --- | --- |
| Bladder | 0.842 (0.696–0.926) | 0.909 (0.722–0.975) | 0.867 (0.758–0.931) |
| Left femoral head | 0.667 (0.391–0.862) | 0.933 (0.821–0.977) | 0.877 (0.768–0.939) |
| Right femoral head | 0.800 (0.548–0.930) | 0.872 (0.733–0.944) | 0.852 (0.734–0.923) |
| Prostate | 0.891 (0.770–0.953) | 0.714 (0.454–0.883) | 0.850 (0.739–0.919) |
| Rectum | 0.976 (0.874–0.996) | 0.579 (0.363–0.769) | 0.850 (0.739–0.919) |

### Qualitative Evaluation of LLM Outputs

Table 6 summarizes the organ-specific results of the qualitative evaluation of the LLM outputs. The overall mean score ± standard deviation (SD) was 1.70 ± 0.48. Among the 291 evaluated LLM outputs, 155 achieved a maximum score of 2 across all criteria, whereas only one output received a score of 0 across all criteria. Representative examples are illustrated in Figure 4.

**Figure 4.**
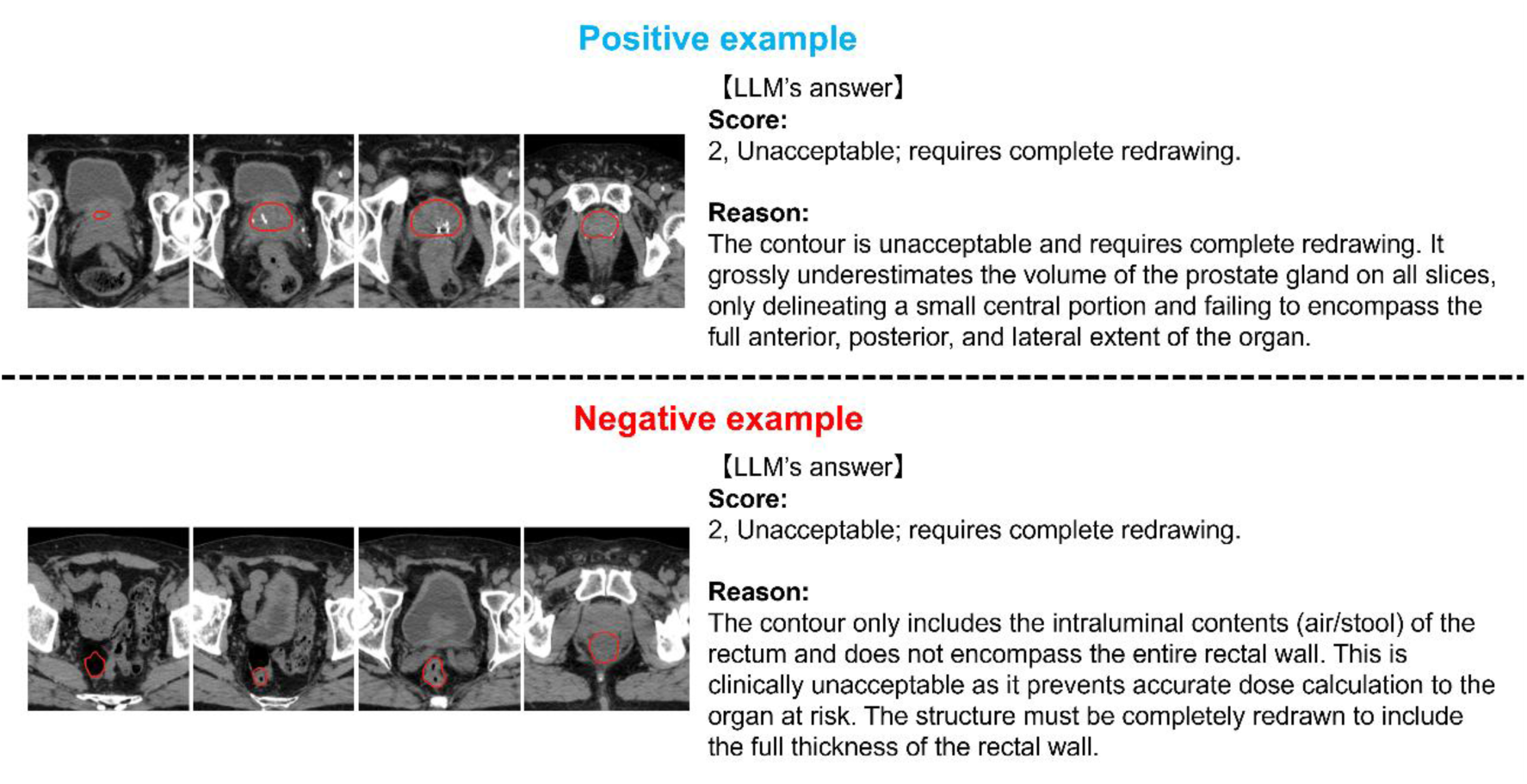
Specific examples of organ evaluation outputs generated by the LLM. The top row demonstrates a positive example, while the bottom row shows a negative example. In the positive example, the LLM accurately points out that the auto-contouring of the prostate (red line) is circumferentially small, achieving a perfect score in the output evaluation by the radiation oncologists. The LLM scored this case as 2 points (requires complete redrawing), which was consistent with the visual assessment by the radiation oncologists. In contrast, in the negative example, the LLM is distracted by gas in the sigmoid colon and fabricates (i.e., hallucinates) an auto-contouring error in the lower rectum. This case was given a score of 0 in the output evaluation by the radiation oncologists. LLM: Large Language Model

**Table 6.** Mean visual evaluation scores of large language model performance by organ.

|  | Accuracy of Error Detection | Absence of Hallucination | Clinical Relevance | Anatomical Understanding |
| --- | --- | --- | --- | --- |
| Bladder | 1.52 ± 0.54 | 1.70 ± 0.50 | 1.73 ± 0.48 | 1.77 ± 0.43 |
| Left femoral head | 1.65 ± 0.48 | 1.67 ± 0.48 | 1.72 ± 0.45 | 1.86 ± 0.35 |
| Right femoral head | 1.63 ± 0.49 | 1.65 ± 0.52 | 1.74 ± 0.44 | 1.87 ± 0.34 |
| Prostate | 1.50 ± 0.50 | 1.78 ± 0.42 | 1.80 ± 0.40 | 1.83 ± 0.38 |
| Rectum | 1.42 ± 0.62 | 1.70 ± 0.50 | 1.60 ± 0.59 | 1.82 ± 0.43 |

(mean ± SD)

## Discussion

In recent years, while the use of LLMs as evaluators, often referred to as “LLM-as-a-Judge”, has been extensively studied in the field of natural language processing, its application to the medical imaging domain remains limited^31^. In this study, we developed and evaluated the utility of the LAQUA system, which utilizes an LLM to perform fully automated QA for AC in radiotherapy. Consequently, the evaluation scores generated by the LLM demonstrated a strong correlation (Spearman’s rank correlation coefficient: 0.733–0.794) with the expert assessments provided by two experienced radiation oncologists. Furthermore, the qualitative evaluation of the LLM outputs showed positive results, with an overall mean score of 1.70 ± 0.48. Over half of the outputs (155/291) achieved a perfect score of 2 across all criteria, while only one received a 0. These findings indicate the potential of the LAQUA system to serve as an effective primary screening tool in the AC quality assurance process.

Conventional QA for AC has predominantly relied on methods that detect delineation errors by extracting geometric features or perform verification using independently trained AC models. However, it has long been recognized that geometric metrics, typically represented by the vDSC, do not necessarily correlate with clinical significance^32^. The primary advantage of the LAQUA system in this study lies in its capability to provide specific feedback in natural language using an LLM. Beyond mere geometric agreement or simple pass/fail judgments, it can explicitly specify error locations, such as “the cranial boundary of the prostate is overestimated” or “the anterior wall of the rectum is missing due to the influence of rectal contents.” Although a recent preprint has explored an LLM-based evaluation approach for medical image segmentation, to the best of our knowledge, there are no reports investigating an LLM-based QA workflow specifically targeting clinically available AC software^33^. Automation bias toward AC outputs carries the risk of causing critical incidents. By providing descriptive text detailing the specific errors that require correction, this system can effectively guide the evaluator’s attention and is expected to mitigate the risk of oversight.

In the qualitative evaluation of the LLM judgments, the LAQUA system achieved a Likert scale score of 1.70 ± 0.48. While this demonstrated high overall validity, it also highlighted clear weaknesses. Representative examples of the qualitative evaluation are shown in Figure 4. In the positive example, the LLM accurately pinpointed typical error locations using natural language, demonstrating strong agreement with the rationales provided by human experts for their scoring. Conversely, in the negative case, the LLM was distracted by gas in the cranial portion; despite adequate contouring in the caudal region, it inappropriately concluded that the entire rectum was poorly contoured and hallucinated a non-essential claim that this would affect dose calculation. These errors were attributed to both the misinterpretation and hallucinations, suggesting that they stem from limitations in the LLM’s image recognition capabilities and its reliance solely on pre-trained, general-purpose knowledge^34,35^. Previous studies have noted a degradation in LLM performance within highly specialized domains^19,36^. It is highly probable that the Gemini model used in this study either lacked specific knowledge of contouring atlas guidelines in radiotherapy, or conflated them with other generalized information. To address this issue, incorporating a Retrieval-Augmented Generation (RAG) system could be an effective solution. Previous reports have demonstrated that integrating reliable external data sources for dynamic reference by the LLM can significantly enhance task accuracy^37–39^. Supplying the LAQUA system with contouring guidelines and detailed anatomical knowledge as an external knowledge base could mitigate errors and achieve further improvements in accuracy. This remains a crucial subject for future research.

It is important to note that the LAQUA system is not intended to completely replace the quality assurance process for AC. Indeed, the sensitivities of the LAQUA system were 0.791 (OncoStudio) to 0.933 (syngo.via) (when a score of ≥ 4 was defined as “adequate”), indicating that its detection capability is not flawless. The primary role of this system is to facilitate a human-in-the-loop framework, wherein human experts review the AI-generated auto-contours and make the final clinical decisions^40–43^. By verbalizing and presenting the specific regions requiring modification to human evaluators, the LAQUA system can alleviate expert workload while simultaneously mitigating automation bias, thereby reducing the likelihood of critical AC errors being overlooked.

This study has several limitations. First, as it utilized a limited, publicly available male pelvic dataset, the robustness of the system against real-world data remains uncertain. Second, because Gemini 2.5 Pro did not support the direct input of DICOM files, this study supplied the 3D CT data to the LLM in the form of 2D PDFs. Consequently, this rendering process altered the original image matrix size, and may have led to the loss of inter-slice continuity as well as contrast information dependent on specific Window Level and Window Width settings, which potentially affected the evaluations.

## Conclusion

In conclusion, this study developed and evaluated the LAQUA system, a fully automated QA system for AC powered by an LLM. Overall, the system’s evaluations demonstrated a strong correlation with expert assessments, suggesting its potential utility as an effective primary screening tool.

## Data Availability

All data produced in the present study are available upon reasonable request to the authors.

https://www.cancerimagingarchive.net/collection/prostate-anatomical-edge-cases/

## Acknowledgements

RatoGuide was provided by AiRato Inc. under a collaborative research agreement. OncoStudio was provided free of charge by Oncosoft Inc. for this study.

We partially used Gemini for English language editing, but the final text was fully reviewed and corrected by the authors.

As all patient data were obtained from a publicly available, de-identified open dataset, ethical approval by an institutional review board was not required for this study.

## Notes

Conflict of interest: Kadoya was a board member and, along with Tozuka, held stock in AiRato Inc., while Onishi and Jingu received grants from AiRato Inc.

### Competing Interest Statement

Kadoya was a board member and, along with Tozuka, held stock in AiRato Inc., while Onishi and Jingu received grants from AiRato Inc.

### Funding Statement

This study was funded by AiRato Inc.

### Author Declarations

As all patient data were obtained from a publicly available, de-identified open dataset (https://www.cancerimagingarchive.net/collection/prostate-anatomical-edge-cases/), ethical approval by an institutional review board was not required for this study.

